# Gender disparities in access to care for time-sensitive conditions during COVID-19 pandemic in Chile

**DOI:** 10.1101/2020.09.11.20192880

**Authors:** Jorge Pacheco, Francisca Crispi, Tania Alfaro, María Soledad Martínez, Cristóbal Cuadrado

## Abstract

**Introduction:** During the COVID-19 pandemic reduction on the utilisation of healthcare services are reported in different contexts. Nevertheless, studies have not explored specifically gender disparities on access to healthcare.

**Aim:** To evaluate disparities in access to care in Chile during the COVID-19 pandemic from a gender-based perspective.

**Methods:** We conducted a quasi-experimental design using a difference-in-difference approach. We compared the number of weekly confirmed cases of a set of oncologic and cardiovascular time-sensitive conditions at a national level. We defined weeks 12 to 26 as an intervention period and the actual year as a treatment group. We selected this period because preventive interventions, such as school closures or teleworking, were implemented at this point. To test heterogeneity by sex, we included an interaction term between difference-in-difference estimator and sex.

**Results:** A sizable reduction in access to care for patients with time-sensitivity conditions was observed for oncologic (IRR 0·56; 95% CI 0·50-0·63) and cardiovascular diseases (IRR 0·64; 95% CI 0·62-0·66). Greater reduction occurred in women compared to men across diseases groups, particularly marked on myocardial infarction (0·89; 95% CI 0·85-0·93), stroke (IRR 0·88 IC95% 0·82-0·93), and colorectal cancer (IRR 0·79; 95% CI 0·69-0·91). Compared to men, a greater absolute reduction in women for oncologic diseases (782; 95% CI 704-859) than cardiovascular diseases (172; 95% CI 170-174) occurred over 14 weeks.

**Conclusion:** We confirmed a large drop in new diagnosis for time-sensitive conditions during the COVID-19 pandemic in Chile. This reduction was greater for women. Our findings should alert policy-makers about the urgent need to integrate a gender perspective into the pandemic response and its aftermath.

**Research in context:** We searched PubMed, Google Scholar and medRxiv using the search terms "Health Services” AND “Access* AND “gender” AND (“pandemics” OR “COVID” OR “SARS-CoV2”) on the title and abstract for research published in 2020, with no language restrictions. Reports of a decrease in healthcare access were common during the pandemic for cardiovascular and oncologic diseases in various countries. Only three studies explored gender differences in access to healthcare for time-sensitive conditions. These studies did not find a differential impact between genders. None of these studies were conducted in settings with higher levels of gender inequalities such as Latin America.

**Added value of this study:** To our knowledge, this is the first study in Latin America that explores gender differences in access to care during the COVID-19 pandemic. We confirmed a large decrease in medical diagnosis in women compared to men for a broad group of time-sensitive cardiovascular and oncologic diseases. We used a comprehensive and reliable national database to test our hypothesis. The effect was evident in conditions with different etiological mechanisms, so it is highly implausible to explain our finding through biological causes. Gender norms and hierarchies better explain this wide effect. An increase in care workload due to school closure and aggravation of gender bias due to scarcity could explain this reduced healthcare utilisation in women during the pandemic.

**Implications of all the available evidence:** Our findings should alert policy-makers about the urgent need to integrate a gender perspective on the current outbreak response. If school closure has a role in the observed differential effect, increasing healthcare services availability will not shorten disparities between sexes. Services provision should enhance access during COVID-19 pandemic, especially for women who are raising children or have other caregiver responsibilities.

## Introduction

The COVID-19 pandemic reduced the utilisation of health care services, similarly to the phenomena reported in previous epidemic outbreaks, like SARS,^1^ MERS,^2^ and Ebola.^3^ In the current pandemic, studies have shown a decrease in the frequency of different interventions like surgeries (electives or not) and hospital admissions, including specific time-sensitive conditions, such as acute coronary syndrome,^4,5^ myocardial infarction,^6,7^ stroke,^7-11^ and cancer.^12-17^

Although it has been largely studied that gender impacts access to healthcare, gender and sex differences in access to healthcare have been scarcely examined during the COVID-19 pandemic. While most studies have not explored heterogeneity of the reduction by gender,^4,9-15^ some studies that examine differences by sex on acute coronary syndrome ^5,6^ and stroke,^8^ have not found relevant disparities. Only one study was done in Latin America and did not explore gender differences.^11^ To our knowledge, a single research explored access differences in cancer care by sex during a pandemic. The authors did not identify any relevant differences, although the more considerable decrease was for breast cancer.^17^ Gender has been proposed as a structural determinant of health, as gender norms shape social stratification, health-related exposures and behaviours, healthcare access, health systems, and health research.^18^ Nevertheless, the response to outbreaks has been usually devoid of a gender perspective, limiting the effectiveness of the public health response^19,20^. Sex refers to the biological differences between men, women and intersex. These biological differences between sexes can produce differential vulnerability to infectious diseases. For example, for COVID-19, some sex-specific mechanisms have been suggested as a relevant factor for worse disease outcomes in men, such as hormone-regulated expression of genes or immune response.^21^

Gender norms and stratification could influence social and economic outcomes, which in turn could impact access to health care (Figure 1). First, mounting evidence has demonstrated that school closure and mandatory confinement has increased caregiving responsibilities in families, which traditionally fall on women, producing significant disruption in their daily lives compared with men. Second, as there is a general reduction in the availability of health services, gender bias that usually affects access for women, especially to cardiovascular diseases, may increase.^21^ Finally, during the pandemic, employment was impacted, and many people suffered income reduction. As women are overrepresented in informal jobs, they experienced higher unemployment rates and a more significant reduction in working hours and salaries compared with men during the pandemic in different contexts.^22,23^ Also, COVID-19 has increased levels of gender violence, and reproductive health is usually not prioritised during emergencies,^21^ potentially reducing access to relevant diagnostic services such as smear test for cervical cancer.

**Figure 1.**
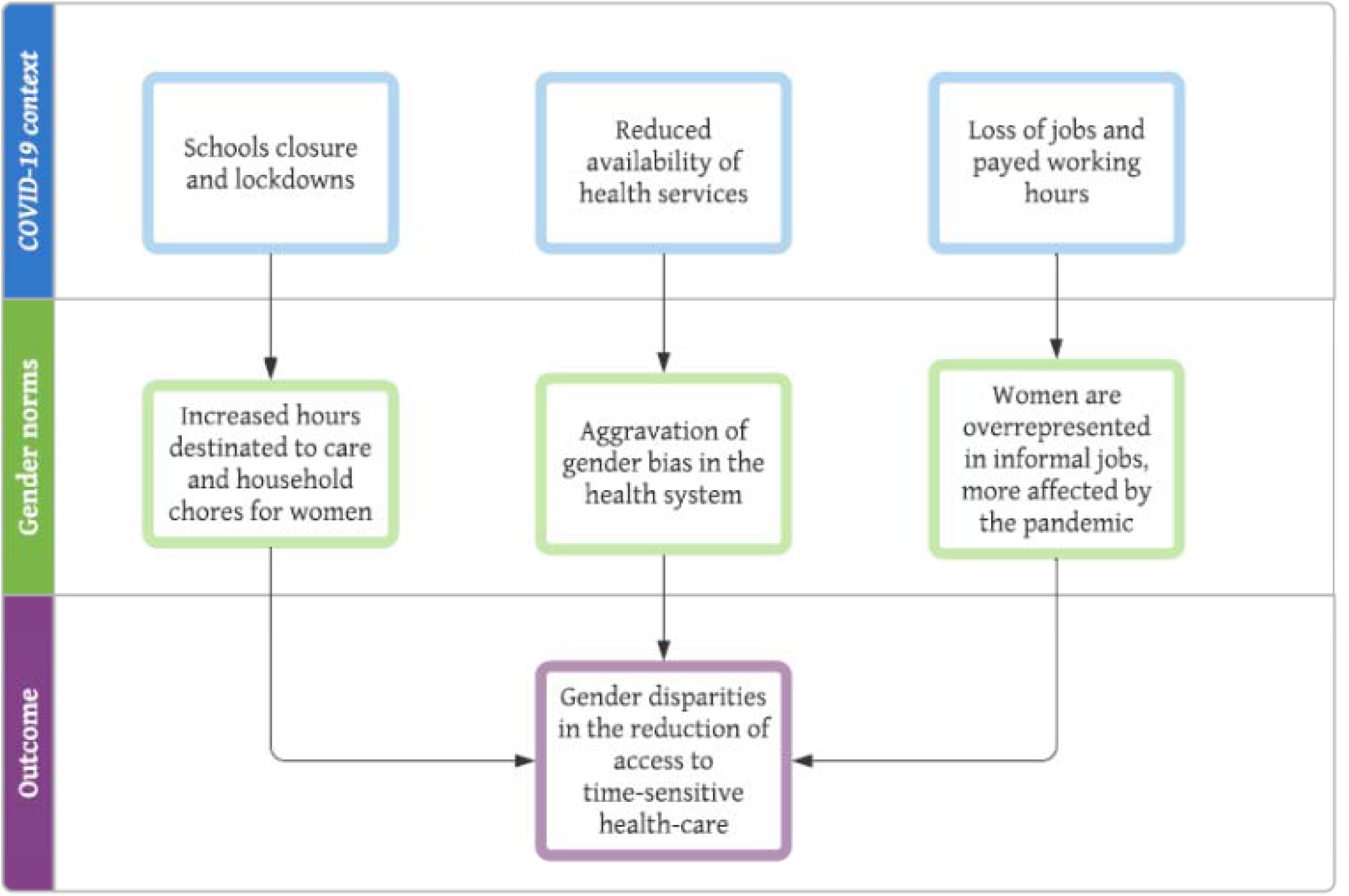
Gender impact on healthcare access during COVID-19. Source: Authors contribution

The sex-gender system is difficult to separate for analysis. In this study, we state that the measured differences in access to healthcare are explained mainly to gender norms. First, because the role of gender in access to healthcare has been previously studied as a relevant factor.^18^ Second, because it seems less plausible that the variations between men and women in the utilisation before and after the pandemic are due to biological characteristics. Therefore, hereafter the paper refers to “gender” for the studied categories of female and male.

During a pandemic, people have massively delayed their consultation due to fear of contagion and reduced availability of medical services. Additionally, we pose that women were differentially affected in the outbreak response due to gender-roles. This study aims to evaluate disparities in access to care in Chile during the COVID-19 pandemic from a gender-based perspective. We focus on severe and time-sensitive group conditions (cardiovascular diseases and cancer) with guaranteed access in the context of the Chilean health system. As observed in other countries, we hypothesised a large drop in both group conditions diagnosis,^4-17^ but with a more significant decrease in women.

## Methods

### Study setting

In 2005 Chile implemented a Health Reform which included the National Explicit Health Guarantees Regime (“AUGE”), a set of guarantees aimed to ensure access to timely (opportunity guarantee), affordable, and quality services for people of both insurance systems predominant in Chile (public, National Health Fund - FONASA-, and private, ISAPRES), for 56 health conditions, which have been amplified to 80 nowadays.^24^ During the current pandemic, the obligation for FONASA and ISAPRES to comply with the Explicit Guarantee of opportunity established for the health problems was suspended for up to one month since the 8th of April, except for severe conditions included in this study such as acute myocardial infarction, stroke, and cancers.

### Data sources

We obtained data from the National Health Fund (Fondo Nacional de Salud - FONASA) which finances all public hospitals in Chile and provides health coverage to nearly 15 million inhabitants (75% of the Chilean population). We selected a set of nine time-sensitive conditions included in the National Explicit Health Guarantees Regime (“AUGE”): two acute cardiovascular diseases (stroke and myocardial infarction) and seven cancers (gastric cancer, colorectal cancer, lymphoma, leukaemia, cervical cancer, breast cancer, and testis cancer).

The attending physician registers every public insured patient with a medical diagnosis of these conditions as a confirmed case. National clinical guidelines standardise the diagnostic process for each disease, reducing practice variation and improving reporting quality. A confirmed case report is mandatory by law for healthcare providers. A description of case definitions included in National Clinical Guidelines is available in the Supplemental material (Table S1).

### Analysis

We conducted a quasi-experimental design using a difference-in-difference method.^25^ Due to the count nature of the data (number of cases diagnosed per week), we fitted generalised linear models with a Negative Binomial distribution. We compared the number of weekly confirmed cases from epidemiological week 1 to 26 for the years 2017 to 2020. We defined week 12 to 26 as an intervention period and year 2020 as a treatment group, because on week 12 started most of the public health interventions implemented during the pandemic, including school closures and remote working recommendations. In that period started a process of cessation of electives surgeries and centralisation of acute beds by the Ministry of Health too. Interventions and dates details are available in the Supplemental material (Tabla S2).

The outcome variable was the number of confirmed cases for the following diseases: stroke (includes transient ischemic attack), myocardial infarction, all cardiovascular diseases (stroke plus myocardial infarction), gastric cancer, colorectal cancer, lymphoma, leukaemia, cervical cancer (includes dysplasia), breast cancer, testis cancer, and all cancers. The independent variables were difference-in-differences estimators and age (20 to 29 years, 30 to 39 years, 40 to 49 years, 50 to 59 years, 60 to 69 years, 70 to 79 years, 80 years, and more). We used as exposure or offset variable the number of persons covered by the national public health insurance by age group to adjust for size and demographic composition. Also, we adjusted for weekly and yearly seasonal trends. The model was defined as:

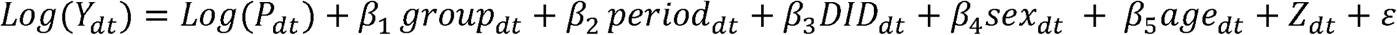

With *Y_dt_* the number of confirmed cases of disease *d* in week *t*, *P_dt_* the population (number) of public health beneficiaries by age-group and sex, *β*_1_represents the effect of being in the treated (year 2020) versus the control group (years 2017-2019), *β*_2_represents the effect pandemic period (week 12 and onwards) versus pre-pandemic period (week 1 to 11), *β*_3_represents the difference-in-difference estimator, *β*_4_captures the effect of sex, *β*_5_adjust for the effect of age. *Z* is a vector of adjustment co-variables, including a set of weekly and yearly time dummy variables, and *ε* is the error term.

To test for heterogeneity by sex, our main effect of interest, we included an interaction term between DID and sex (Model 2). As a sensitivity analysis, we fitted unadjusted and adjusted models for all cancers, after excluding sex-specific cancers (breast cancer, cervical cancer, and testicular).

We reported the mean and standard deviation for descriptive analysis and incidence rate ratios (IRR) and absolute effects (counts) with 95% confidence intervals for regressions models. We used STATA 16.0 for analyses and R 4.0.2 for graphs.

### Ethics

Since this study used secondary data from publicly available sources collected by the Ministry of Health, which are registered anonymously, we did not require institutional review board approval.

## Results

We analysed a total of 214 465 cardiovascular events (54 308 strokes and 160 157 myocardial infarction) and 91 655 cancer diagnosis (8 199 gastric cancers, 15 500 colorectal cancers, 3 506 lymphomas, 1 745 leukaemia, 28 865 cervical cancers, 24 760 breast cancers, and 1 725 testis cancers) during the study period (Table 1).

**Table 1.** Weekly number of confirmed cases by disease and year.

|  | 2017 | 2018 | 2019 | 2020 |
| --- | --- | --- | --- | --- |
|  | Mean (SD) | Mean (SD) | Mean (SD) | Mean (SD) |
| Stroke (includes transient ischemic attack) | 493.3 (32.6) | 583.5 (33.2) | 580.0 (38.4) | 476.9 (101.4) |
| Myocardial infarction | 1552.2 (87.8) | 1607.4 (65.2) | 1698.2 (78.3) | 1302.1 (355.6) |
| Gastric cancer | 154.4 (25.5) | 166.5 (25.6) | 169.9 (31.2) | 107.4 (67.1) |
| Colorectal cancer | 103.5 (11.5) | 145.7 (20.6) | 205.8 (30.3) | 141.2 (87.9) |
| Lymphoma | 31.6 (6.6) | 32.9 (6.9) | 40.2 (7.1) | 30.2 (9.3) |
| Leukaemia | 17.0 (4.0) | 16.5 (4.3) | 19.3 (5.4) | 14.3 (7.0) |
| Cervical cancer (includes dysplasia) | 263.2 (41.5) | 295.3 (40.7) | 316.8 (37.3) | 234.9 (131.6) |
| Breast cancer | 226.8 (24.3) | 256.0 (31.1) | 274.9 (36.4) | 194.6 (99.6) |
| Testicular cancer | 16.5 (5.1) | 16.4 (4.0) | 18.5 (4.4) | 14.9 (6.6) |

Compared to previous years (2017-2019), after week 12 (March 15, 2020) an immediate downward trend in the number of events was confirmed for all diseases (figure 1). In the oncologic diseases group, a smaller decrease occurred for lymphoma and leukaemia (figure 1). In the cardiovascular diseases group, we observe more substantial reductions for myocardial infarction compared to stroke. Previous to the observed downward, trends were parallel for all studied diseases (figure 1). When analysed by sex, women showed a greater impact on their access compared with men for both diseases groups during the study period (figure 2).

**Figure 2.**
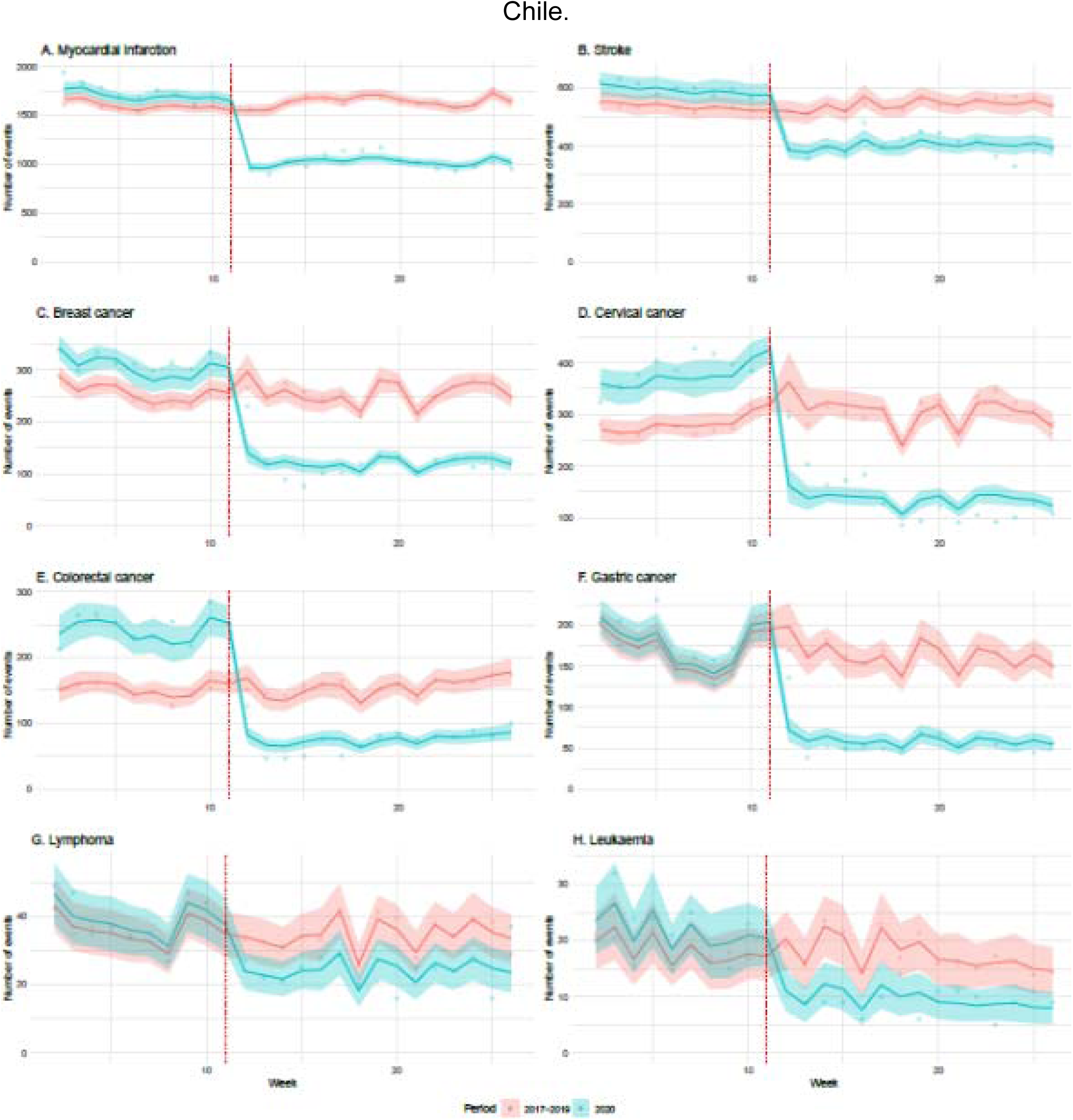
Trends in confirmed cases for cardiovascular and oncologic diseases. 2017-2020, Chile. Figure 2: Points represent the average number of events (new cases diagnosed) per week for each disease during the first 26 weeks of the year. Solid lines are the point estimate for the fitted model. Coloured areas around the lines are the 95% confidence intervals for the fitted model. In red, the cases observed in years 2017-2019 (used as a control group). In green, the number of patients diagnosed in 2020 (affected by the COVID-19 pandemic). The vertical dotted line represents the starting week of the first population-level interventions for COVID-19 in Chile (week 11). Each panel (A to H) illustrates the trend of a different disease.

**Figure 3.**
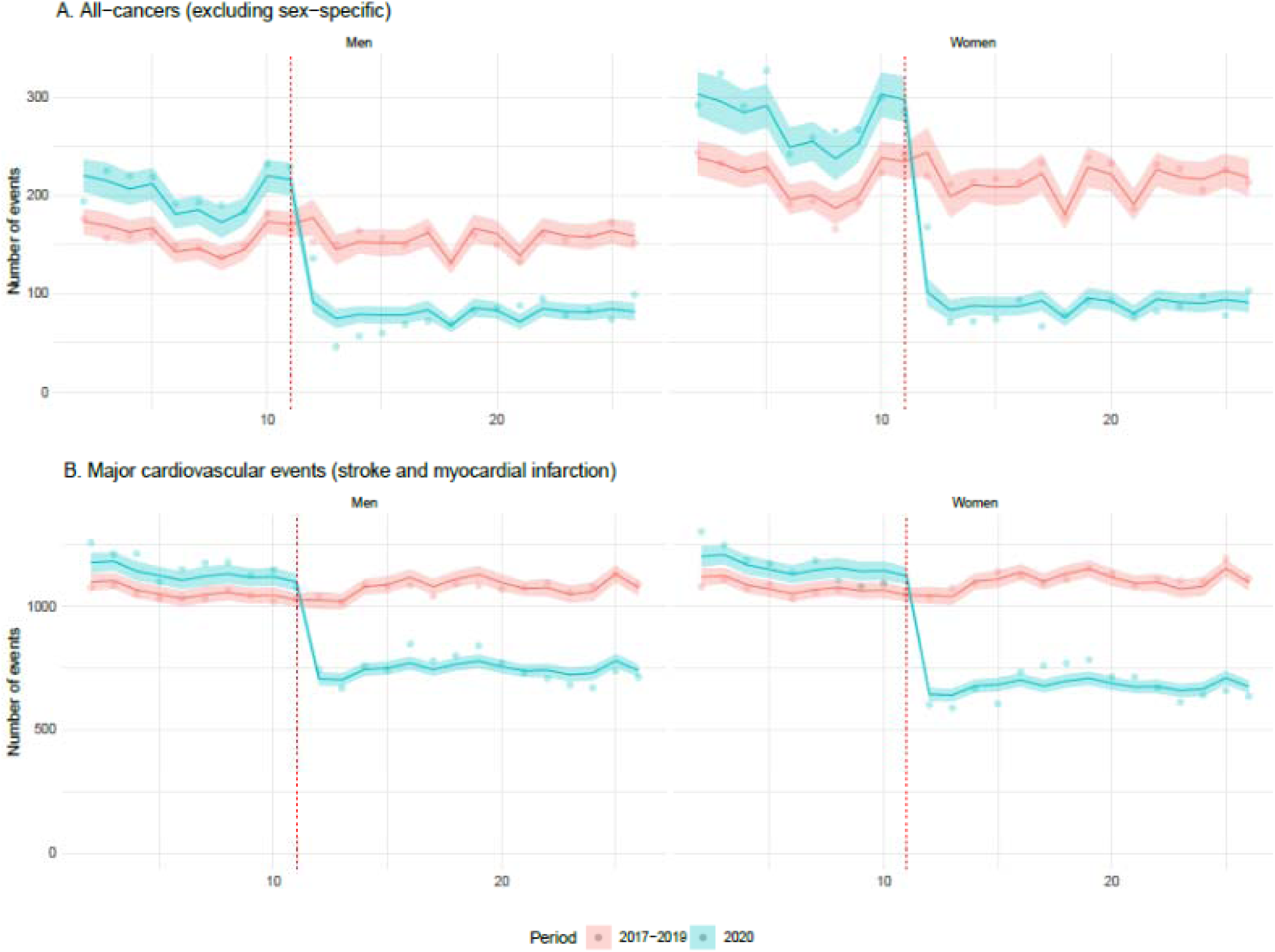
Trends in confirmed cases for cardiovascular and oncologic diseases group (excluding sex-specific cancers) during the study period. Figure 3: Points represent the average number of events (new cases diagnosed) per week for all-cancers (exhibit A) and cardiovascular events (exhibit B) during the first 26 weeks of the year. Cancers exclude sex-specific conditions such as breast, cervical or testicular cancer to facilitate comparisons between sexes. Cardiovascular events include stroke and myocardial infarction. Right panels present data for women; left panels data for men. Solid lines are the point estimate for the fitted model. Coloured areas around the lines are the 95% confidence intervals for the fitted model. In red, the cases observed in years 2017-2019 (used as a control group). In green, the number of patients diagnosed in 2020 (affected by the COVID-19 pandemic). The vertical dotted line represents the starting week of the first population-level interventions for COVID-19 in Chile (week 11).

In our model, we confirmed a larger reduction for cancer conditions (IRR 0·56; 95% CI 0·50-0·63) compared to the cardiovascular events (IRR 0·64; 95% CI 0·62-0·66). Among cancer conditions, a greater reduction was observed in colorectal cancer (IRR 0·35; 95% CI 0·31-0·39), cervical cancer (IRR 0·34; 95% CI 0·30-0·37), gastric cancer (IRR 0·38; 95% CI 0·33-0·44) and breast cancer (IRR 0·40; 95% CI 0·37-0·43). In the cardiovascular group, the most affected condition was myocardial infarction (IRR 0·62; 95% CI 0·60-0·64) (Table 2).

**Table 2.** Incidence Rate Ratio for confirmed cases during pandemic period (week 12-16) †.

|  | DID estimator<br>IRR (95% CI) | Interaction between sex and DID estimator<br>IRR (95% CI) |
| --- | --- | --- |
| All cardiovascular diseases | 0.64 (0.62-0.66) | 0.89 (0.86-0.92) |
| Stroke (includes transient ischemic attack) | 0.72 (0.68-0.76) | 0.88 (0.82-0.93) |
| Myocardial infarction | 0.62 (0.60-0.64) | 0.89 (0.85-0.93) |
| All cancer | 0.56 (0.50-0.63) | 0.76 (0.67-0.86) |
| All cancer (excluding sex specific cancer) | 0.41 (0.37-0.45) | 0.81 (0.72-0.91) |
| Gastric cancer | 0.38 (0.33-0.44) | 0.86 (0.72-1.03) |
| Colorectal cancer | 0.35 (0.31-0.39) | 0.79 (0.69-0.91) |
| Lymphoma | 0.70 (0.57-0.85) | 0.87 (0.70-1.08) |
| Leukaemia | 0.49 (0.36-0.66) | 0.88 (0.62-1.24) |
| Cervical cancer (includes dysplasia) | 0.34 (0.30-0.37) | .. |
| Breast cancer | 0.40 (0.37-0.43) | .. |
| Testicular cancer | 0.50 (0.38-0.65) | .. |

† Difference-in-difference model adjusted by sex, age, population size, population age-sex structure, and seasonality (week and year). The model includes an interaction term between DID estimator and sex. Complete models are available in Supplemental material (Table S3-S5)

A differential impact, with larger effects on women than men, was observed across cardiovascular and oncological diseases. For the former, a 11% (IRR 0·89; 95% CI 0·86-0·92) additional reduction in access for major cardiovascular events in women compared with men was observable. For the latter, a further 24% reduction (0·76; 95% CI 0·67-0·86) in access to newly diagnosed cancers among females compared with males was evident in the pandemic period. In the sensitivity analysis, differences between sexes in the cancer group persisted after excluding sex-specific cancers such as breast, cervical, and testicular cancers. In this analysis, a bigger impact on access was observable among women (IRR 0·81 IC95% 0·72-0·91) (Table 2).

When analysed by specific cardiovascular diseases, a greater decrease in women than men took place for stroke (IRR 0·88; 95% CI 0·82-0·93) and myocardial infarction (0·89; 95% CI 0·85-0·93). When analysed by specific oncologic diseases, a greater decrease in women than men occurred for colorectal cancer (IRR 0·79; 95% CI 0·69-0·91). Also, a greater reduction on cervical cancer (IRR 0·33; 95% CI 0·30-0·37) and breast cancer (IRR 0·37; 95% CI 0·37-0·43) compared to testicular cancer (IRR 05; 95% CI 0·38-0·65) was observed (Table 2).

To make sense of these findings, we also present absolute effects sizes. Greater absolute effects in access for women compared to men occurred in almost all conditions. An excess impact in women compared to men was observed for oncologic (782; 95% CI 704-859) and cardiovascular diseases (172; 95% CI 170-174) during the 14 weeks of the pandemic included in the study period. In the sensitivity analysis, differences between sexes persisted but were smaller (109; 95% CI 106-112). When analysed by specific diseases, we found sizable differences in access for women compared to men for myocardial infarction (120 95% CI 52-68), gastric cancer (73; 95% CI 32-41) and colorectal cancer (52 95% CI 23-28) (Table 3).

**Table 3.** Absolute reduction in confirmed cases during the pandemic period (week 12-16)

|  | Men<br>Count (95%CI) | Women<br>Count (95%CI) | Excess impact on woman<br>Count (95%CI) |
| --- | --- | --- | --- |
| All cardiovascular diseases | 774 (726-822) | 946 (896-996) | 172 (170-174) |
| Stroke (includes transient ischemic attack) | 152 (129-175) | 211 (188-234) | 59 (59-59) |
| Myocardial infarction | 621 (581-661) | 741 (698-783) | 120 (117-123) |
| All cancer | 156 (128-183) | 937 (883-1 042) | 782 (704-859) |
| All cancer (excluding sex specific cancer) | 197 (181-212) | 305 (287-324) | 109 (106-112) |
| Gastric cancer | 115 (102-128) | 188 (172-204) | 73 (69-76) |
| Colorectal cancer | 95 (87-102) | 146 (137-156) | 52 (50-54) |
| Lymphoma | 11 (6-16) | 14 (9-19) | 3 (3-3) |
| Leukaemia | 9 (6-12) | 11 (8-14) | 2 (2-2) |
| Cervical cancer (includes dysplasia) | .. | 420 (394-446) | .. |
| Breast cancer | .. | 318 (298-339) | .. |
| Testicular cancer | 18 (13-24) | .. | .. |

## Discussion

In our study, we confirmed a large drop in the medical diagnosis for time-sensitive conditions during the COVID-19 pandemic in Chile. This decrease was more significant for oncologic than cardiovascular diseases. Also, we confirmed our hypothesis of gender disparities in medical diagnosis. A large group of time-sensitive conditions were affected by this differential effect, even though healthcare access for these conditions is guaranteed by law to everyone. This finding is worrisome because delaying care for these severe conditions can lead to long-term disability and - eventually - premature death.

Gender is a structural determinant of health^18^ and, usually, is not prioritised during an outbreak response.^19,20^ Through different mechanisms, gender could affect healthcare access in a pandemic. Nevertheless, differential access by gender has not been adequately studied during this current outbreak. Only a few studies explored heterogeneity by sex in cardiovascular^5,6,8^ and cancer care^17^ without finding any significant effect. None of these studies were done in Latin America.

Because these diseases have different etiological mechanisms, it is highly implausible to explain this finding through biological causes. While a stroke and myocardial infarction could increase after COVID-19 infection,^26,27^ a reduction in the number of cardiovascular events is probably due to decreased access. If men are more prone to COVID-19, ^21^ this could explain, at least partially, that the decline in stroke and myocardial infarction in males could be smaller compared with women. Although, this explanation cannot be given in cancer because these diseases do not share the same causes and acute changes in cancer incidence are unlikely to be attributable to COVID-19 infection. In this setting, a reduced number of newly diagnosed cancers, particularly among women, is a clear marker of reduced access and unmet needs.

Gender norms and hierarchies could explain better this wide effect. Differences in help-seeking behaviour between genders have been commonly described.^29^ On average, Chilean women used more healthcare services than men.^30^ During COVID-19, school closure to control disease transmission has a differential effect on women because they provide most of the informal care in families.^19,20^ A greater differential effect was observed on diseases (colorectal, cervical, gastric, and breast cancer) that require scheduled appointments for testing. Added to income reduction due to work hours decrease^22^ and unemployment,^23^ an increase in domestic workload can decrease women’s time availability and reduce healthcare demand during a pandemic.

From the supply side, gender biases^18^ - heuristics based on gender stereotypes - have been associated with delayed access to cardiovascular treatment in women.^28^ Potentially, these biases could aggravate during the pandemic. Service offering reduction could promote severe disease prioritisation by medical teams, which could unintentionally reduce healthcare access for women. For instance, in the context of scarcity, the treatment for only the standard male could be prioritised.

This study has several limitations. First, we use administrative data, which might be subject to underreporting during the pandemic. Nevertheless, confirmed case reports have been mandatory for healthcare providers since 2004. Moreover, they are used for health claim payments in the Chilean health system, therefore making it less likely that reduced reporting could explain the observed effect. Second, due to the data codification, this study only considers two categories for sex and gender (female and male). This dichotomy excludes a spectrum of gender identities and the intersex population.^18^ Future studies must explore differential effects on health care accessibility during pandemics for a broader gender classification. Third, we cannot rule out residual confounding in the context of observational data. Nevertheless, due to the characteristic of the exposure of interest (the pandemic) is unlikely that better data could be obtained using alternative sources or study designs. We controlled confirmed cases by population and age in our models and included seasonal adjustments by week and year to control for unobserved time-specific confounding factors. The use of previous year trends as a control for the same observational units allows adjustment for confounding, but since no parallel control group was available adjustment for other time-variant effects concomitants to the pandemic was not feasible.

As strengths, this is the first study from Latin America that explores access by sex to medical diagnosis during the COVID-19 pandemic. To test our hypothesis, we used a rich, comprehensive, and reliable national database where cases were defined based on standardised diagnostic processes. We select a variety of severe time-sensitive conditions to avoid generalisation based on anecdotal evidence. Moreover, we tested different models, maintaining our conclusions unchanged.

As previous researchers have posed, ^19,20^ our findings should alert policy-makers about the urgent need to integrate a gender perspective into an outbreak response. If school closure has a role in the observed differential effect, increasing healthcare services availability will not shorten disparities between sexes. Services provision should enhance access, especially for women who are raising children or have other caregiver responsibilities and reduce economic barriers. Also, health professionals should be aware of this situation and encouraged through clinical guidelines to reduce current gender bias in their clinical practice. Future research must evaluate the consequences of access reductions on disability and premature death. The observed effect occurred in a set of severe time-sensitive conditions where care delays could worsen prognosis. Additionally, we need to know the causes, which could be informed through surveys and innovative ways to provide care for these diseases during the actual pandemic.

In conclusion, as previous studies have shown,^4-17^ we confirmed a large drop in medical diagnosis for cardiovascular and oncologic conditions in Chile during COVID-19 pandemic. Additionally, we demonstrate that women were far more affected compared to men. This differential effect by gender was observed for a broad group of time-sensitive conditions. Because these conditions have different etiological mechanisms, biological causes are unlikely to explain our findings. Gender norms and hierarchies define better this differential effect. Emergent healthcare barriers, such as an increase in care work due to school closure, aggravation of gender bias, and income reduction could decrease healthcare access in women during pandemics and potentially cause long-term disability and premature death in them. Our study should alert policymakers and put women’s access to healthcare as a top global health priority during this pandemic.

## Data Availability

Data and codes are publicly available.

https://github.com/CoV-IMPACT-C/gender-impact-access-covid

## Contributors

JP did literature research, collected data, developed the study design, analyzed data and drafted the manuscript. FC did literature research, design figures, interpreted data and drafted the manuscript. TA did literature research, analyzed data, interpreted data and drafted the manuscript. MSM interpreted data and critically revised the manuscript. CC developed the study design, interpreted data, design graphs, and drafted the manuscript.

## Acknowledgments

The authors thanks FONASA for their support during doing this research. This work was funded by the National Agency for Research and Development (ANID), Scholarship program, DOCTORADO BECAS CHILE 2020 - 21200241 and COVID research fund ANID-COVID0960.

## References

1. Chang HJ, Huang N, Lee CH, Hsu YJ, Hsieh CJ, Chou YJ. The impact of the SARS epidemic on the utilization of medical services: SARS and the fear of SARS. American Journal of Public Health. 2004: 94(4): 562-564. DOI: 10.2105/ajph.94.4.562

2. Lee SY, Khang YH, Lim HK. Impact of the 2015 Middle East Respiratory Syndrome Outbreak on Emergency Care Utilization and Mortality in South Korea. Yonsei Med J. 2019;60(8):796–803. doi:10.3349/ymj.2019.60.8.796

3. Ribacke KJ, Saulnier DD, Eriksson A, von Schreeb J. Effect of the West Africa Ebola Virus Disease on health-care utilization. A systematic review. Frontiers in public health. 2016 Oct 10;4:222. doi: 10.3389/fpubh.2016.00222. eCollection 2016

4. De Filippo O, D’Ascenzo F, Angelini F et al. Reduced rate of hospital admissions for ACS during COVID-19 outbreak in Northern Italy. NEJM 2020 Apr 28; NEJMc2009166. doi: 10.1056/NEJMc2009166.

5. Mafham MM, Spata E, Goldacre R et al. COVID-19 pandemic and admission rates for and management of acute coronary syndromes in England. The Lancet. 14 de julio del 2020. DOI: https://doi.org/10.1016/S0140-6736(20)31356-8

6. Solomon MD, McNulty EJ, Rana JS et al. The Covid-19 pandemic and the incidence of acute myocardial infarction. NEJM 2020. May 19, DOI: 10.1056/NEJMc2015630

7. Baum A, Schwartz MD. Admissions to Veteran Affairs Hospitals for Emergency Conditions during the COVID-19 pandemic. JAMA. Published online June 05, 2020. doi:10.1001/jama.2020.99729.

8. Rudilosso S, Laredo C, Vera V. et al. Acute stroke care is at risk in the era of COVID-19. Experience at a comprehensive stroke center in Barcelona. Stroke 2020, 51: 1991-1995. https://doi.org/10.1161/STROKEAHA.120.030329

9. Hoyer C, Ebert A, Hagen B et al. Acute stroke in times of the COVID-19 pandemic. A Multicenter study. Stroke 2020, 51: 2224-2227. https://doi.org/10.1161/STROKEAHA.120.030395

10. Zhao J, Li H, Kung D et al. Impact of the COVID-19 epidemic on stroke care and potential solutions. Stroke 2020, 51: 1996-2001. https://doi.org/10.1161/STROKEAHA.120.030225

11. Diegoli H, Magalhaes PS, Martins SC et al. Decrease in Hospital Admissions for Transient Ischemic Attack, Mild, and Moderate Stroke During the COVID-19 Era. Stroke 2020, 51(8):2315-2321. https://doi.org/10.1161/STROKEAHA.120.030481

12. Dinmohamed AG, Visser O, Verhoeven RH et al. Fewer cancer diagnosis during the COVID-19 epidemic in the Netherlands. The Lancet Oncology 2020. 21(6): 750-751. DQI:https://doi.org/10.1016/S1470-2045(20)30265-5

13. Rutter MD, Brookes M, Lee TJ, et al. Impact of the COVID-19 pandemic on UK endoscopic activity and cancer detection: a National Endoscopy Database AnalysisGut Published Online First: 20 July 2020. doi: 10.1136/gutjnl-2020-322179

14. Guven DC, Aktas BY, Aksun MS, et al. COVID-19 pandemic: changes in cancer admissions. BMJ Supportive & Palliative Care Published Online First: 14 July 2020. doi: 10.1136/bmjspcare-2020-002468

15. Zadnik, V., Mihor, A., Tomsic, S., Zagar, T., Bric, N., Lokar, K., & Oblak, I. (2020). Impact of COVID-19 on cancer diagnosis and management in Slovenia - preliminary results, Radiology and Oncology, 54(3), 329-334. doi: https://doi.org/10.2478/raon-2020-0048

16. Filipe D, van Deukeren D, Kip M et al. Impact of the COVID-19 pandemic on surgical breast cancer care in the Netherlands: a multicentre retrospective cohort study. Clinical Breast Cancer 2020. https://doi.org/10.1016/j.clbc.2020.08.002.

17. Kaufman HW, Chen Z, Niles J, Fesko Y. Changes in the Number of US Patients With Newly Identified Cancer Before and During the Coronavirus Disease 2019 (COVID-19) Pandemic. JAMA Netw Open. 2020;3(8):e2017267. Published 2020 Aug 3. doi:10.1001/jamanetworkopen.2020.17267

18. Heise L, Greene ME, Opper N, et al. Gender inequality and restrictive gender norms: framing the challenges to health. Lancet. 2019;393(10189):2440–2454. doi:10.1016/S0140-6736(19)30652-X

19. Smith J. Overcoming the “tyranny of the urgent”: integrating gender into disease outbreak preparedness and response. Gender & Development 2020 27(2):355-369. DOI: 10.1080/13552074.2019.1615288

20. Wenham C, Smith J, Morgan R. COVID-19: the gendered impacts of the outbreak, The Lancet 2020. 395(10227):846-848 DQI:https://doi.orq/10.1016/S0140-6736(20)30526-2

21. Gebhard C, Regitz-Zagrosek V, Neuhauser HK, Morgan R, Klein SL. Impact of sex and gender on COVID-19 outcomes in Europe. Biol Sex Differ. 2020;11(1):29. Published 2020 May 25. doi:10.1186/s13293-020-00304-9

22. Collins, C, Landivar, LC, Ruppanner, L, Scarborough, WJ. COVID-19 and the gender gap in work hours. Gender Work Organ. 2020; 1-12. https://doi.org/10.1111/gwao.12506

23. Landivar LC, Ruppanner L, Scarborough WJ, Collins C. Early Signs Indicate That COVID-19 Is Exacerbating Gender Inequality in the Labor Force. Socius: Sociological Research for a Dynamic World. 2020. 6:1-3. https://doi.org/10.1177/2378023120947997

24. Frenz P, Delgado I, Kaufman JS, Harper S. Achieving effective universal health coverage with equity: evidence from Chile, Health Policy and Planning, Volume 29, Issue 6, September 2014, Pages 717-731, https://doi.org/10.1093/heapol/czt054

25. Abadie A. Difference-in-difference estimators. En Durlauf SN, Blume LE (Eds). Microeconomics (2010). The New Palgrave Economics Collection. Palgrave MacMillan, London.

26. Merkler AE, Parikh NS, Mir S, et al. Risk of Ischemic Stroke in Patients With Coronavirus Disease 2019 (COVID-19) vs Patients With Influenza. JAMA Neurol. Published online July 02, 2020. doi:10.1001/jamaneurol.2020.2730

27. Long B, Brady WJ, Koyfman A et al. Cardiovascular complications in COVID-19. The American Journal of Emergency Medicine. 2020. 38(7):1504-1507

28. Ahmed Haider, Susan Bengs, Judy Luu, Elena Osto, Jolanta M Siller-Matula, Taulant Muka, Catherine Gebhard, Sex and gender in cardiovascular medicine: presentation and outcomes of acute coronary syndrome, European Heart Journal, Volume 41, Issue 13, 1 April 2020, Pages 1328-1336, https://doi.org/10.1093/eurheartj/ehz898

29. Hunt K., Adamson J., Galdas P. (2010) Gender and Help-seeking: Towards Gender-comparative Studies. In: Kuhlmann E., Annandale E. (eds) The Palgrave Handbook of Gender and Healthcare. Palgrave Macmillan, London. https://doi.org/10.1057/9780230290334_13

30. Vega M, Jeanette, Bedregal G, Paula, Jadue H, Liliana, Delgado B, Iris. (2003). Equidad de género en el acceso a la atención de salud en Chile. Revista médica de Chile, 131(6), 669-678. https://dx.doi.org/10.4067/S0034-98872003000600012

